# Structural, Functional and Cognitive Validation of a Deep-Learning MCI-to-AD Conversion Model in OASIS-3

**DOI:** 10.64898/2026.09.05.26362318

**Authors:** Sara Fin, Alireza Moayedikia

## Abstract

**Background:** Deep-learning models predict mild cognitive impairment (MCI)-to-Alzheimer’s disease (AD) conversion from structural MRI with high accuracy. However, they are typically validated within a single cohort and judged on discrimination alone. Whether their risk scores are biologically grounded, and whether they generalise to independent data, remains unclear.

**Methods:** We applied the ADNI-trained Temporal Adaptive Fusion Network (TAF-Net), without retraining, to 101 MCI participants from OASIS-3 (25 converters, 76 stable) and tested whether its conversion-risk scores were grounded in independent structural, functional, and cognitive markers of AD. Regional atrophy rates were derived from longitudinal FreeSurfer, cognition from longitudinal MMSE and CDR-Sum-of-Boxes, and baseline resting-state functional connectivity from fMRIPrep, in structural and functional subsamples of 41 and 40 participants. Associations used rank-based statistics with false-discovery-rate correction.

**Results:** Higher TAF-Net risk tracked faster atrophy in medial-temporal AD-signature regions but not in AD-spared cortex — an anatomically specific coupling that survived adjustment for global atrophy. In external validation, risk discriminated converters (AUC = 0.72), comparable to native atrophy and strongly concordant with it; atrophy statistically accounted for the model’s prognostic signal. Discrimination transferred but calibration did not: the two lowest tertiles of risk were assigned near-zero probability yet converted at 15%. Risk also tracked the multi-year rate of cognitive decline and, at baseline, was associated with reduced within-network functional-connectivity integrity, concentrated in salience and default-mode hubs; longitudinal functional analyses were underpowered.

**Conclusions:** A conversion model trained on one cohort produced risk scores that, in an independent cohort, were grounded in the structural, functional, and cognitive hallmarks of Alzheimer’s disease — supporting biological validity and external generalisation. The score, however, largely re-expresses the neurodegenerative substrate captured by structural atrophy. Rank ordering transferred across cohorts but absolute risk did not, so the score requires recalibration before its values can be interpreted as individual probabilities.

## 1. Introduction

Alzheimer’s disease (AD) is the most common cause of dementia and develops over a prolonged preclinical and prodromal course, during which amyloid deposition precedes tau pathology and the accompanying neuronal loss that correlates most closely with cognitive decline [1, 2]. Mild cognitive impairment (MCI) is the symptomatic pre-dementia stage and a critical window for intervention, yet its outcomes are heterogeneous: a substantial proportion of patients progress to AD while many remain stable or revert to normal cognition [3]. Accurately identifying which individuals with MCI will convert is therefore central both to clinical management and to the efficient design of therapeutic trials, where patient selection is a major determinant of trial feasibility and cost [4].

Structural magnetic resonance imaging (MRI) captures the neurodegeneration that drives conversion. Atrophy in AD follows a stereotyped topography, beginning in the medial temporal lobe — entorhinal cortex, hippocampus, and amygdala — and spreading to temporoparietal association cortex, while primary sensorimotor and visual cortices are relatively spared until late; this pattern mirrors the spread of tau pathology and underpins MRI-based staging and prediction [5, 6, 7]. Rates of change estimated from longitudinal scans capture prognostic information beyond single-timepoint measures [7, 8].

Deep-learning models increasingly predict MCI-to-AD conversion from structural MRI [9, 10, 11]. The Temporal Adaptive Fusion Network (TAF-Net) models paired longitudinal 3D MRI through an adaptive temporal-gating fusion mechanism and, on the Alzheimer’s Disease Neuroimaging Initiative (ADNI), achieves the highest discrimination among the structural-MRI-only methods compared in that study (test AUC = 0.916), approaching multimodal approaches that additionally require positron-emission tomography, cerebrospinal fluid, or genetic data [12]. A related dual-model framework from the same group extends prognostication in AD from baseline cerebrospinal-fluid biomarkers, pairing a probabilistic trajectory network with a deep survival model [13].

Despite strong discrimination metrics, deep-learning conversion models are typically evaluated within a single cohort and judged on predictive accuracy alone. Two questions therefore remain open. First, do such models generalise to independent cohorts acquired on different scanners and populations? Second, and more fundamentally, are their risk scores biologically grounded — that is, do they reflect genuine AD neurodegeneration rather than dataset-specific or non-specific image features? Internal interpretability analyses such as saliency or attention maps [14, 15] are suggestive but cannot by themselves establish grounding. Independent, external validation against established structural, functional, and cognitive markers of AD is needed before such models can be trusted for clinical or trial use [16].

The present study addresses this gap by design. Rather than reporting discrimination alone, we subject a transferred conversion model to multi-modal external biological validation, asking whether a single risk score converges with three independent axes of AD pathophysiology — structural atrophy, cognitive decline, and functional-network integrity — in a cohort never seen during training. Such convergent, cross-modal grounding provides stronger evidence of construct validity than accuracy metrics or single-modality checks, and offers a template for validating imaging-based AI biomarkers more generally.

We apply the ADNI-trained TAF-Net, without any retraining, to an independent OASIS-3 MCI cohort [17], and evaluate five hypotheses: (1) TAF-Net risk correlates with FreeSurfer-measured regional atrophy rates (association); (2) this association is anatomically specific to AD-signature regions and absent in AD-spared cortex (specificity); (3) in the external cohort, TAF-Net risk discriminates converters comparably to structural atrophy and is concordant with it, with atrophy statistically accounting for its prognostic signal (prognostic concordance); (4) TAF-Net risk tracks the multi-year trajectory of cognitive decline (cognitive concordance); and (5) TAF-Net risk is associated with resting-state functional-connectivity integrity (functional concordance).

## 2. Methods

### 2.1 Participants and dataset

Participants were drawn from the Open Access Series of Imaging Studies-3 (OASIS-3) [17]. The analysis cohort comprised 101 individuals with mild cognitive impairment (MCI; global Clinical Dementia Rating [CDR] = 0.5) at the baseline MRI, each with at least two longitudinal T1-weighted (T1w) structural MRI sessions. Participants were classified as converters (progressive MCI, pMCI; n = 25) if they reached global CDR *≥* 1 within 36 months of the baseline MCI scan, and as stable (sMCI; n = 76) otherwise. Nineteen participants contributed more than one eligible scan pair. Because the 36-month conversion window is measured from each pair’s own baseline, conversion status is defined at the level of the scan pair. A participant was classified as a converter if any eligible pair met the criterion, and their risk score was the mean across eligible pairs. Baseline characteristics by group are summarised in Table 1 and compared in Section 3.1. Age was taken from the Uniform Data Set visit closest to the baseline MRI and was entered as a covariate in the cognitive analyses; the structural analyses adjusted for global atrophy and head size but not age (see Section 5).

**Table 1.** Baseline characteristics by outcome group.

| Characteristic | All (n=101) | pMCI (n=25) | sMCI (n=76) | Difference (95% CI) | p |
| --- | --- | --- | --- | --- | --- |
| Sex, M / F / unknown | 58 / 39 / 4 | 17 / 5 / 3 | 41 / 34 / 1 | - | 0.083 |
| Male, % | 60% | 77% | 55% | - | - |
| Baseline MMSE, mean (SD) | 27.2 (2.5) | 26.0 (3.0) | 27.6 (2.2) | -1.6 (-2.9, -0.3) | 0.017 |
| Age at baseline MRI, yr, mean (SD) | 73.6 (7.8) | 72.9 (7.3) | 73.9 (7.9) | -1.0 (-4.4, +2.2) | 0.43 |
| Baseline global CDR | 0.5 | 0.5 | 0.5 | - | - |
| TAF-Net risk, mean (SD) | 0.16 (0.28) | 0.31 (0.37) | 0.11 (0.23) | +0.21 (+0.06, +0.37) | <0.001 |
| median (IQR) | 0.007<br>(0.0005–0.120) | 0.161<br>(0.002–0.699) | 0.004<br>(0.0003–0.056) | – | – |
| Structural pair interval, mo (median) | 29.4 | 25.0 | 35.8 | - | <0.001 |
| Cognitive follow-up, yr (median) | 8.8 | - | - | - | - |

pMCI, progressive (converter) MCI; sMCI, stable MCI; MMSE, Mini-Mental State Examination; CDR, Clinical Dementia Rating; IQR, interquartile range. Dashes mark quantities not reported for categorical variables, for CDR (no variance), or for the structural interval (a by-design difference).

The interval between the paired longitudinal structural scans was a median of 29.4 months overall and was shorter in converters than in stable participants (25.0 vs 35.8 months; Mann-Whitney p < 0.001; Table 1) — an expected consequence of the 36-month conversion definition. Because atrophy was expressed as an annualised rate (SPC/yr), this interval difference does not bias the atrophy comparisons. Cognitive follow-up extended over a median of 8.8 years (Table 1). Baseline resting-state functional MRI was available for a subset of participants; where a second functional scan existed it was separated from baseline by a median of 3.5 years (range 1.6–9.6), did not differ significantly between groups (Mann-Whitney p = 0.20), and was frequently acquired on a different scanner (see Functional MRI processing).

### 2.2 TAF-Net conversion-risk scores

Conversion risk was estimated with the Temporal Adaptive Fusion Network (TAF-Net) [12], a hybrid convolutional-transformer architecture that predicts 3-year MCI-to-AD conversion from paired longitudinal 3D T1w MRI via an adaptive temporal-gating fusion module. TAF-Net was trained exclusively on ADNI [18, 19] under subject-level cross-validation with no data leakage, achieving a held-out ADNI test AUC of 0.916. For the present study the ADNI-trained model was applied to OASIS-3 as a fully external test set; no OASIS-3 data were used in training or tuning. Each participant received a continuous conversion-risk score in [0, 1], averaged across eligible scan pairs for the 19 participants with more than one pair. TAF-Net was trained on scan pairs separated by 6–24 months; in OASIS-3 the pair interval ranged from 0.2 to 85.1 months (median 29.4), so most participants were scored at intervals outside the range on which the model was trained, an aspect of transfer examined in Section 3.4.

### 2.3 Structural MRI processing and atrophy quantification

Cortical surface reconstruction and subcortical segmentation used FreeSurfer 8.2.0 [20, 21]. The longitudinal stream [22, 23] was applied: cross-sectional recon-all for each session, an unbiased within-subject base template, and longitudinal runs for each timepoint. Of the 101 participants, longitudinal FreeSurfer output was completed and quality-passed for 45; four with scan intervals below one year were excluded because annualised change rates are unstable over such intervals, yielding 41 participants (15 pMCI / 26 sMCI) for atrophy analyses (Sections 3.2–3.4). The reduction from 101 reflects processing completeness and pipeline failures, not eligibility; all 101 met imaging inclusion criteria.

For each participant and region, atrophy was quantified as the annualised symmetrised percent change (SPC/yr) between the earliest and latest completed timepoints (negative = tissue loss; positive = expansion, expected for ventricles). SPC is the difference between the two timepoints divided by their mean, expressed as a percentage and annualised by the interval between them; it is the rate measure produced by the FreeSurfer longitudinal stream [22]. Cortical regions (Desikan-Killiany atlas [24]) were summarised by mean cortical thickness, vertex-weighted across hemispheres; subcortical structures and lateral ventricles by summed bilateral volume. Estimated total intracranial volume (eTIV) was recorded for head-size normalisation. For the specificity analysis (Section 3.3), regions were assigned a priori to signature AD-vulnerable, prefrontal, spared AD-resistant, and ventricular tiers, following established AD atrophy topography [6, 7].

### 2.4 Functional MRI processing and connectivity

Resting-state functional MRI was available at baseline for 44 of the MCI participants. Functional images were preprocessed with fMRIPrep 24.1.1 [25], comprising head-motion correction, fieldmap-less susceptibility-distortion correction, boundary-based co-registration, and normalisation to MNI152NLin2009cAsym space at 2-mm resolution, with FreeSurfer surface reconstruction disabled. Time-series were denoised by regressing out 24 motion parameters; white-matter, cerebrospinal-fluid, and global signals; and cosine high-pass terms, with censoring of high-motion volumes (framewise displacement > 0.5 mm) [26]. Benchmarking indicates that strategies of this kind — motion expansion, global-signal regression and censoring — minimise the residual motion–connectivity relationship at the cost of some distance-dependent artifact [27]. Four participants retaining fewer than 30 volumes were excluded, leaving 40 usable baseline scans (15 converters, 25 stable); the number of retained volumes was recorded for each participant and used as a covariate in the sensitivity analysis of Section 3.6. Regional time-series were extracted with the Schaefer 400-parcel/7-network atlas [28, 29] and pairwise correlations were Fisher z-transformed [30, 31]. Within-network connectivity was the mean of region-pair correlations within each of the seven networks; whole-brain global connectivity and a default-mode posterior (PCC/precuneus) sub-measure were computed analogously. A within-network integrity composite was defined as the mean of the seven z-scored network measures, with the first principal component as a confirmatory index.

### 2.5 Cognitive measures

The Mini-Mental State Examination (MMSE, 0–30; higher = better) [32] and the CDR Sum-of-Boxes (CDR-SB, 0–18; higher = worse) [33, 34] were obtained from OASIS-3 Uniform Data Set assessments across all available visits. Baseline cognition was taken from the first assessment with a valid score. For each participant, the rate of cognitive change was estimated as the ordinary-least-squares slope per year across all available visits.

### 2.6 Statistical analysis

Given the right-skewed distribution of TAF-Net risk, rank-based statistics were used throughout. Atrophy correlations (Section 3.2): Spearman and Pearson correlations between TAF-Net risk and regional SPC/yr, and converter-stable comparisons by Mann-Whitney U and Welch t tests with Cohen’s d. Anatomical specificity (Section 3.3): partial Spearman correlation between risk and each region’s atrophy, controlling for global cortical atrophy (and eTIV for volumes), with false-discovery-rate (Benjamini-Hochberg) correction [35] across 25 regions and a signature-versus-spared dissociation tested by Steiger’s Z. Prognostic discrimination (Section 3.4): area under the ROC curve (AUC) with bootstrap 95% confidence intervals (2000 resamples for AUCs and connectivity correlations; 10,000 resamples for the Table 1 group differences) and Mann-Whitney U, and logistic regression of conversion on standardised risk and standardised signature atrophy. Cognitive concordance (Section 3.5): Spearman and age-adjusted partial Spearman correlations (age at the first assessment) with baseline cognition and decline rates, each with 10,000-resample bootstrap 95% CIs; converter-stable differences in decline rate by Mann-Whitney U with bootstrap CIs for the mean difference; and a sensitivity analysis in the 41 imaging participants. Functional concordance (Section 3.6): Spearman correlations between risk and each within-network measure, whole-brain connectivity, and the integrity composite, with FDR across the network measures; converter-stable comparison with bootstrap AUC; an exploratory region-wise analysis (FDR across 400 parcels); and longitudinal linear mixed-effects models (time *×* group) with scanner as a covariate. Calibration and operating characteristics (Section 3.7): observed conversion was compared with mean predicted probability by tertile of risk, with Wilson 95% intervals, and the Brier score was referenced to its no-skill value under the observed base rate. Sensitivity, specificity and predictive values were tabulated at the default threshold of 0.5 and at the Youden-optimal point, and the effect of a simple in-sample logistic recalibration of the score on the Brier score was examined. Discrimination in the full cohort used a bootstrap over participants (2000 resamples) and was additionally examined at the level of individual scan pairs (n = 125), stratified by the interval between the paired scans. Functional associations (Section 3.6) were repeated as partial correlations adjusting for the number of retained volumes. Analyses used Python (SciPy, statsmodels, scikit-learn, nilearn); significance was set at p or q < 0.05.

## 3. Results

### 3.1 Cohort

The 101 MCI participants (25 pMCI, 76 sMCI) all had a TAF-Net risk score and baseline cognitive assessments (Table 1). Converters had lower baseline MMSE than stable participants (26.0 vs 27.6; Mann-Whitney p = 0.017) and, as expected, markedly higher TAF-Net risk (0.31 vs 0.11; p < 0.001). The sex distribution differed at a trend level (pMCI 77% vs sMCI 55% male among participants with recorded sex; Fisher exact p = 0.08), and sex is therefore noted as a potential confounder (Section 5). Age did not differ between groups (mean 72.9 vs 73.9 years at the baseline MRI; Mann-Whitney p = 0.43). Forty-one participants had completed, quality-passed longitudinal FreeSurfer atrophy measures (imaging sample, Sections 3.2–3.4); cognitive analyses (Section 3.5) used the full cohort; baseline resting-state fMRI was available for 40 (Section 3.6). The distribution of individual TAF-Net risk scores was strongly zero-inflated: the median score was 0.007 (IQR 0.0005–0.120) and 52% of participants scored below 0.01, with a thin upper tail reaching 0.96 (Figure 7A). The mean and standard deviation in Table 1 are therefore a poor summary of this distribution, and the median and IQR are reported alongside them; throughout, the score is treated as an ordering of participants rather than a graded probability.

### 3.2 TAF-Net risk correlates with regional atrophy

Across the imaging sample (n = 41), higher TAF-Net risk was associated with faster atrophy in medialtemporal regions characteristic of early AD: amygdala (Spearman *ρ* = -0.58, p < 0.001), hippocampus (*ρ* = -0.48, p = 0.001), and entorhinal cortex (*ρ* = -0.46, p = 0.003), and with faster lateral-ventricular expansion (Pearson r = +0.51, p = 0.001) (Figure 1). The per-region scatter plots show the same relationships at the individual level, with converters concentrated at the high-risk, fast-atrophy end of each distribution (Figure 2A). Whole-brain rate was not significantly associated (*ρ* = -0.18, p = 0.27) (Figures 1, 2A). Converters atrophied approximately twice as fast as stable participants in the hippocampus (-2.89 vs -1.42 %/yr; Mann-Whitney p = 0.002, d = -1.03) and in ventricular expansion (+6.72 vs +3.69 %/yr; p = 0.004, d = +1.16) (Figure 2B).

**Figure 1.**
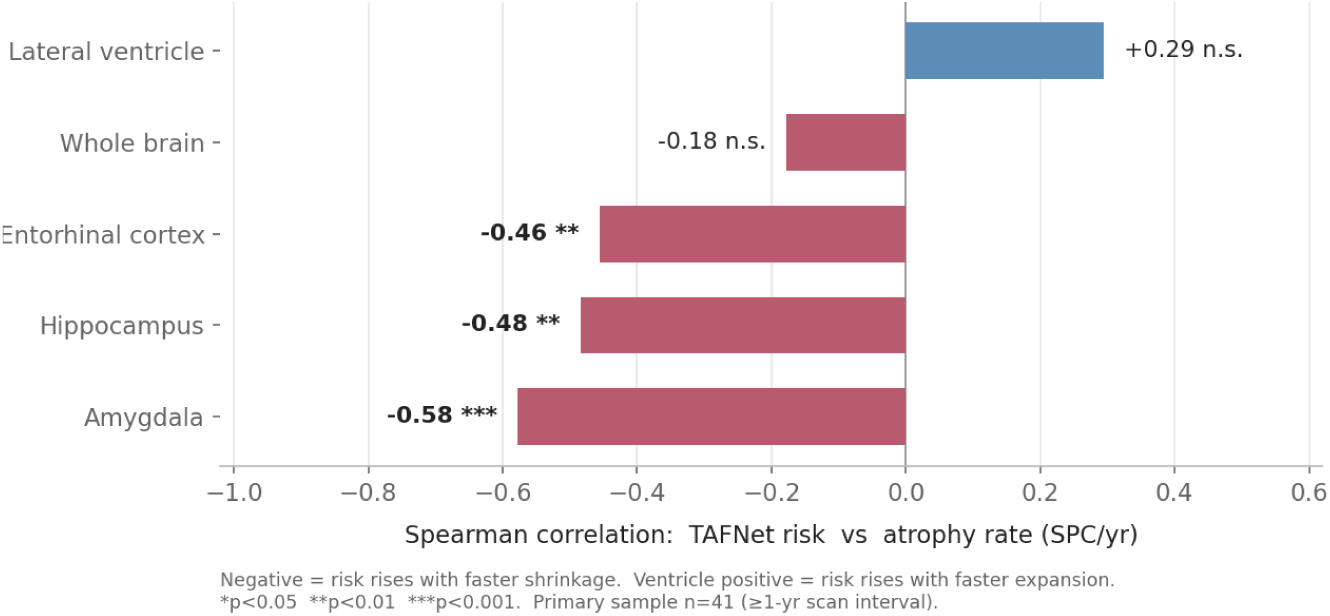
Higher TAF-Net risk tracks faster medial-temporal atrophy Spearman rank correlations between TAF-Net risk and regional annualised atrophy rate (SPC/yr); n = 41. Negative values indicate that higher risk tracks faster shrinkage; the lateral ventricle is positive, denoting faster expansion.

**Figure 2.**
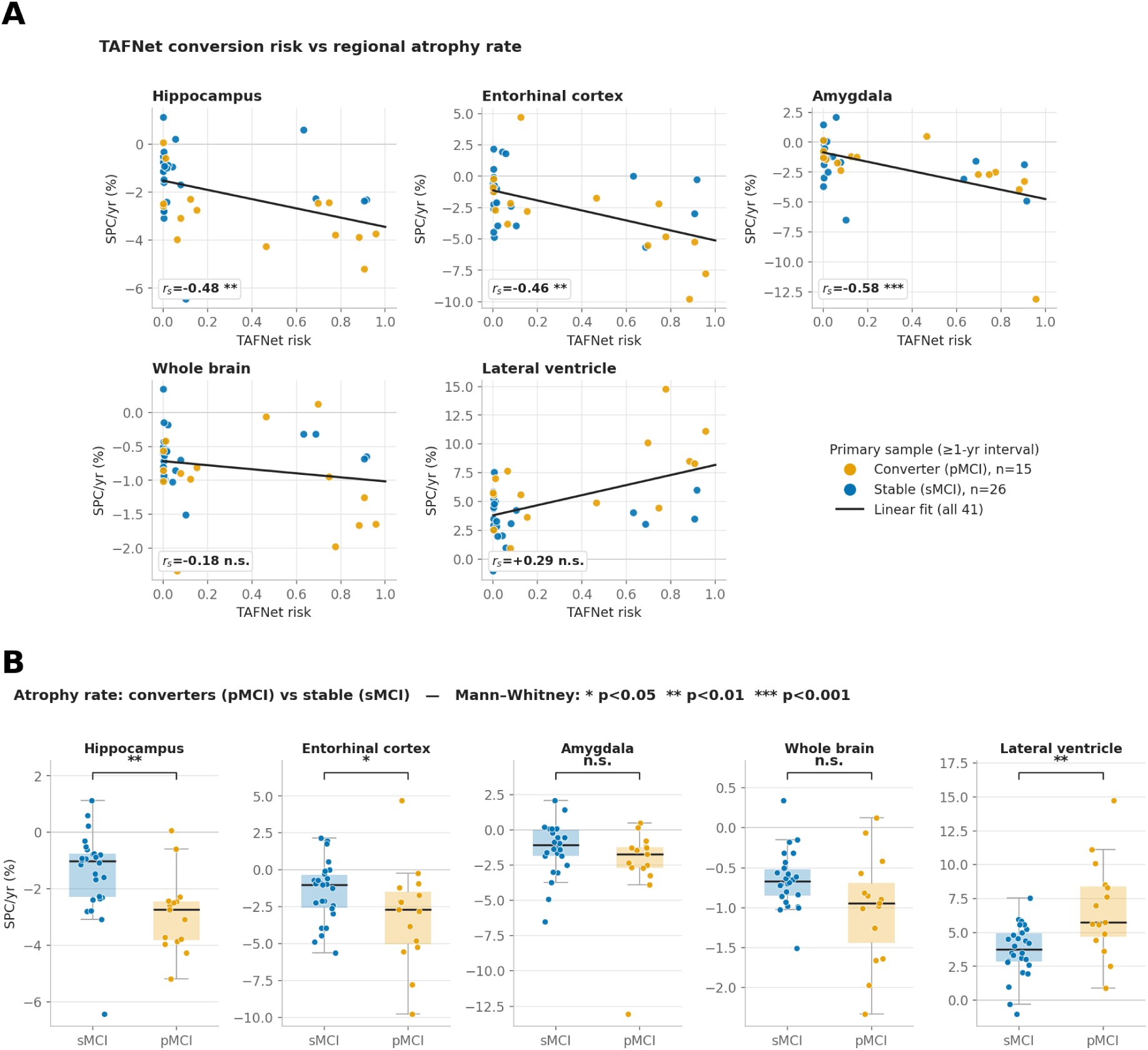
Risk–atrophy associations and converter–stable differences (A) Per-region scatter of TAF-Net risk against annualised atrophy rate with ordinary-least-squares trend; orange, converter; blue, stable. (B) Atrophy rates by group. n = 41 (15 pMCI, 26 sMCI).

### 3.3 The association is anatomically specific

After adjustment for global cortical atrophy (and eTIV for volumes), the risk-atrophy coupling remained significant after FDR correction in six signature regions — entorhinal cortex (partial *ρ* = -0.59, q = 0.002), amygdala (-0.51, q = 0.010), inferior temporal (-0.50, q = 0.010), parahippocampal (-0.48, q = 0.011), middle temporal (-0.46, q = 0.015), and hippocampus (-0.43, q = 0.032) — whereas no spared or prefrontal region survived correction (Table 2). The region-wise pattern is shown for cortical thickness and subcortical volume separately in Figure 3 (left and right panels), where signature regions cluster at negative values and spared regions at zero. A composite dissociation confirmed the pattern: risk was coupled to the signature composite (partial *ρ* = -0.586, p = 0.0001) but not the spared composite (*ρ* = -0.075, p = 0.64); the difference was significant (Steiger Z = -3.23, p = 0.0012; Figure 3). The putamen, coupled at the unadjusted level (*ρ* = -0.34), was non-significant after adjustment (*ρ* = -0.27, q = 0.27).

**Table 2.** Global-atrophy-adjusted partial correlations with regional atrophy (n = 41)

| Region | Tier | Adjusted $\rho$ | q |
| --- | --- | --- | --- |
| Entorhinal cortex | signature | -0.594 | 0.002 |
| Amygdala (volume) | signature | -0.505 | 0.010 |
| Inferior temporal | signature | -0.504 | 0.010 |
| Parahippocampal | signature | -0.483 | 0.011 |
| Middle temporal | signature | -0.463 | 0.015 |
| Hippocampus (volume) | signature | -0.426 | 0.032 |
| Signature composite | - | -0.586 | $p=0.0001$ |
| Spared composite | - | -0.075 | $p=0.64$ |
| Signature vs spared (Steiger) | - | $Z=-3.23$ | $p=0.0012$ |

Spared and prefrontal regions were non-significant and are not shown. q, false-discovery-rate-corrected p.

**Figure 3.**
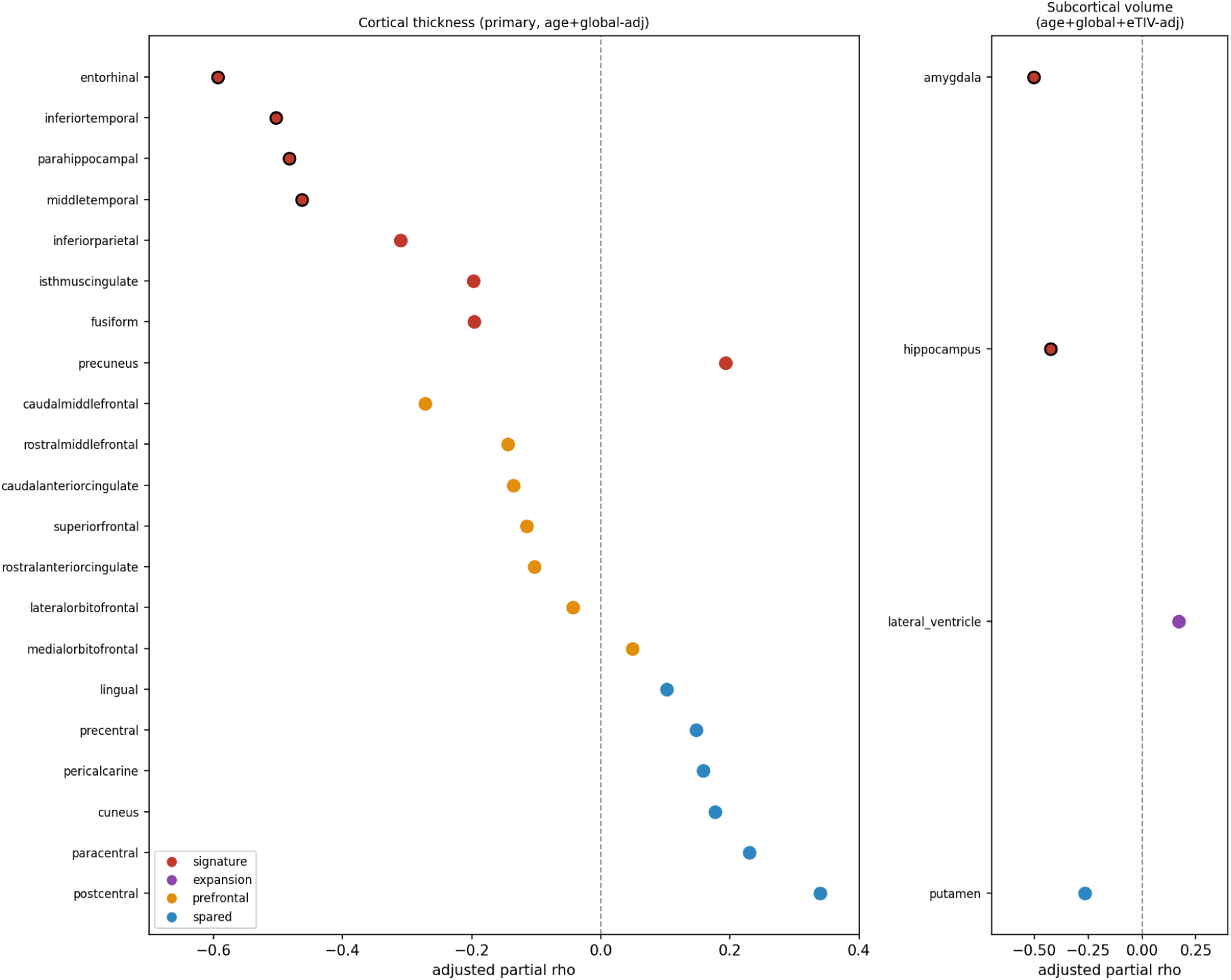
TAF-Net risk couples selectively to AD-signature atrophy Region-specific partial Spearman correlations with atrophy, adjusted for global cortical atrophy and, for volumes, eTIV. Cortical thickness, left panel; subcortical volume, right panel. Filled markers survive Benjamini–Hochberg FDR across 25 regions (q *<* 0.05). Red, signature regions; blue, spared.

### 3.4 External prognostic concordance

Applied to OASIS-3 as an external cohort, TAF-Net risk discriminated converters from stable participants across the full cohort (AUC = 0.72, 95% CI 0.59–0.84; Mann-Whitney p = 0.0009; n = 101; bootstrap over participants; Figure 8A). In the imaging subsample in which native atrophy is also available, discrimination was comparable (AUC = 0.71, 95% CI 0.53–0.88, p = 0.026; n = 41). The comparisons that follow are made within that subsample, since they require FreeSurfer-derived atrophy. Native FreeSurfer atrophy performed similarly — hippocampal atrophy AUC = 0.80 (0.63–0.95) and signature-composite atrophy AUC = 0.79 (0.60–0.93) — with overlapping confidence intervals (Figure 4, left). Converters showed both higher risk and faster signature atrophy than stable participants (Figure 4, right). Risk and signature atrophy were strongly concordant (Spearman *ρ* = -0.59, p = 0.0001). In separate logistic models, each predictor was significant (risk *β* = 0.75, p = 0.030; atrophy *β* = 1.70, p = 0.008). In the joint model, atrophy remained significant (*β* = 1.59, p = 0.018) whereas risk did not (*β* = 0.27, p = 0.53), and the combined model (AUC = 0.81) barely exceeded atrophy alone (0.79) (Figure 4, left). Structural atrophy thus statistically accounted for the model’s prognostic signal.

**Figure 4.**
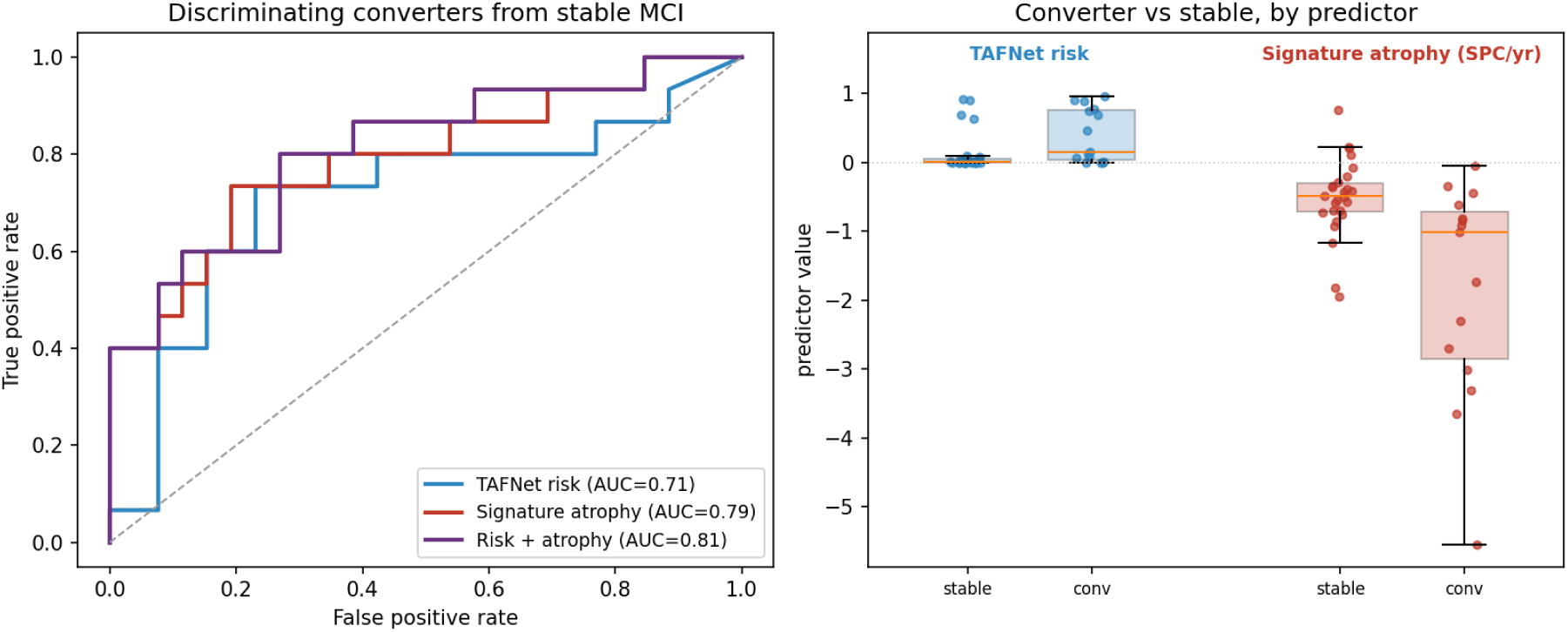
TAF-Net risk discriminates converters concordantly with atrophy (Left) ROC curves for TAF-Net risk, signature-atrophy rate and their combination, with 2000-sample bootstrap confidence intervals. (Right) Risk and signature-atrophy rate by group. n = 41.

At the default threshold of 0.5 the transferred score was markedly conservative: sensitivity 0.32 at specificity 0.89, with a positive predictive value of 0.50 and a negative predictive value of 0.80 (Table 3). The Youden-optimal threshold in this cohort was far lower, at 0.03, where sensitivity rose to 0.76 at a specificity of 0.72 (Figure 8A) — the default operating point is far from optimal when the score is transferred to a new cohort.

**Table 3.** Operating characteristics of the transferred risk score in the full cohort (n = 101, 25 converters)

| Threshold | Sensitivity | Specificity | PPV | NPV | TP | FP | FN | TN |
| --- | --- | --- | --- | --- | --- | --- | --- | --- |
| 0.50 (default) | 0.32 | 0.89 | 0.50 | 0.80 | 8 | 8 | 17 | 68 |
| 0.10 | 0.52 | 0.80 | 0.46 | 0.84 | 13 | 15 | 12 | 61 |
| 0.03 (Youden) | 0.76 | 0.72 | 0.47 | 0.90 | 19 | 21 | 6 | 55 |

Risk is the participant-level score (mean across eligible pairs). PPV, positive predictive value; NPV, negative predictive value; TP, FP, FN, TN, true and false positives and negatives. The Youden-optimal threshold maximises sensitivity plus specificity minus one and was derived in this cohort, so its performance is optimistic.

Discrimination was also examined at the level of individual scan pairs, by the interval between the paired scans (Figure 8B). Within the 6–24-month window on which TAF-Net was trained, AUC was 0.74 (28 pairs, 9 converters); in the 24–60-month stratum, which contains most of the cohort (76 of 125 pairs), it was 0.69. The under-6-month stratum (9 pairs, 4 converters) is too small to be informative, and the over-60-month stratum contains no converters. Ninety-seven of 125 pairs (78%) fall outside the training window, and performance across them degraded modestly rather than collapsing.

### 3.5 Cognitive concordance

In the full cohort (n = 101; median follow-up 8.8 years), higher TAF-Net risk was associated with worse baseline cognition on the MMSE (*ρ* = -0.24, 95% CI -0.41 to -0.05, p = 0.015; age-adjusted *ρ* = -0.22, 95% CI -0.41 to -0.02, p = 0.026) but not with baseline CDR-SB (*ρ* = +0.12, 95% CI -0.08 to +0.31, p = 0.22), as expected in a cohort selected for a uniform global CDR of 0.5. Because worse cognition corresponds to lower MMSE but higher CDR-SB scores, these associations are negative for MMSE and positive for CDR-SB. The longitudinal measures showed stronger coupling: higher risk predicted a faster annual loss of MMSE points (*ρ* = -0.28, 95% CI -0.46 to -0.08, p = 0.005; age-adjusted -0.27, 95% CI -0.46 to -0.07) and, most strongly, a faster annual accumulation of CDR-SB impairment (*ρ* = +0.38, 95% CI +0.19 to +0.54, p = 0.0001; age-adjusted +0.37, 95% CI +0.17 to +0.54) (Figure 5A). Figure 5B summarises these four associations with their 95% confidence intervals, before and after adjustment for age. Both decline-rate intervals excluded zero and lay further from zero than the baseline associations, indicating that the risk score is more informative about the trajectory of cognitive decline than about cognitive status at a single time point. Age adjustment changed none of the estimates materially (Figure 5B, purple), so the associations are not attributable to age differences among participants.

**Figure 5.**
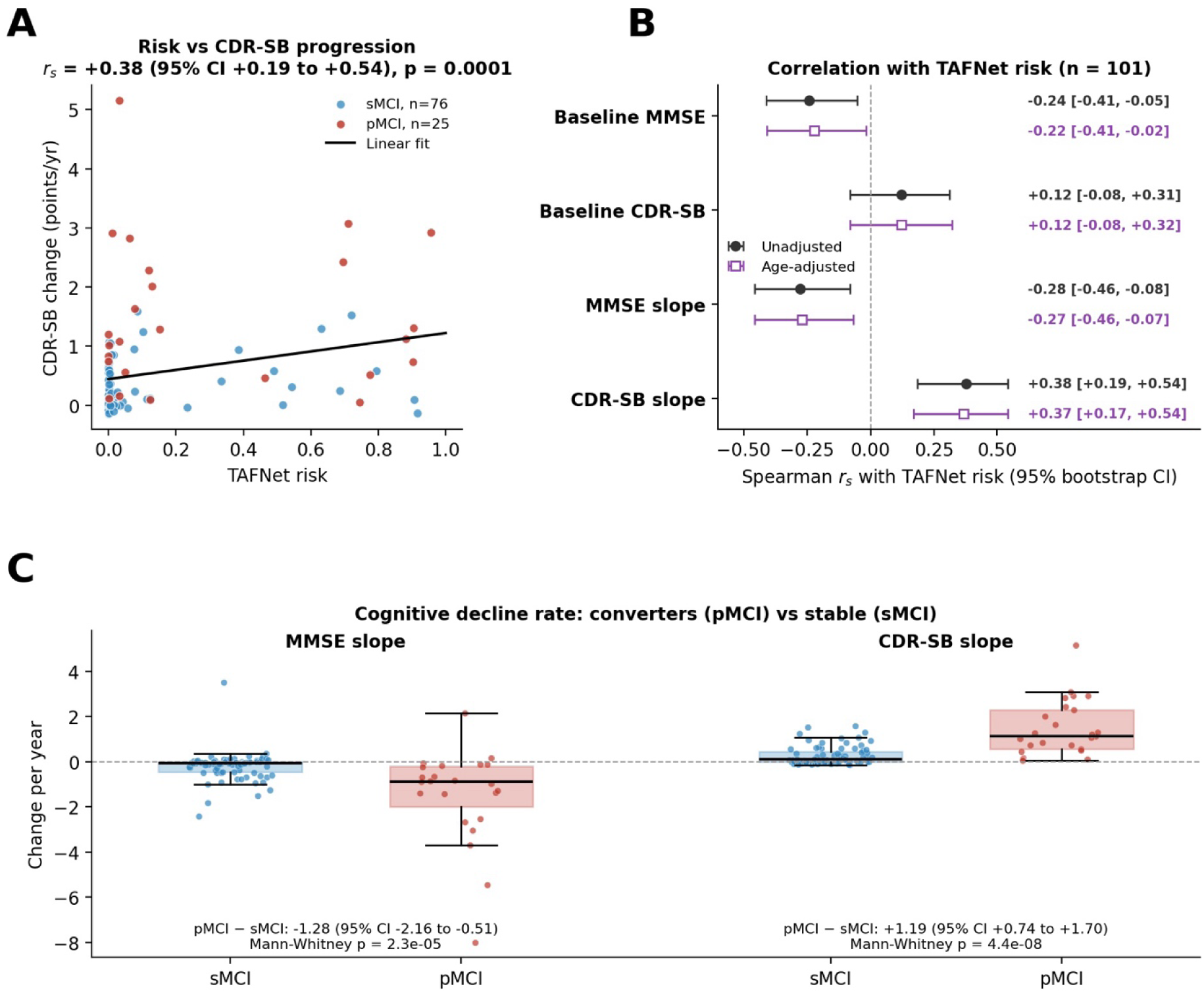
TAF-Net risk tracks the rate of cognitive decline (A) Risk against CDR-SB progression rate; red, converter; blue, stable; dashed line, least-squares fit. (B) Spearman correlations with baseline cognition and decline rates, unadjusted (black) and age-adjusted (purple), with 10,000-resample bootstrap confidence intervals. (C) Decline slopes by group; box, median and interquartile range; points, individual participants.

Converters declined markedly faster than stable participants: MMSE -1.49 vs -0.21 points/yr (difference -1.28, 95% CI -2.16 to -0.51) and CDR-SB +1.46 vs +0.27 per year (difference +1.19, 95% CI +0.74 to +1.70); both Mann-Whitney p < 0.0001 (Figure 5C). A sensitivity analysis in the 41 imaging participants preserved the result (CDR-SB progression *ρ* = +0.33, p = 0.035; MMSE decline *ρ* = -0.29, p = 0.066).

### 3.6 Functional concordance

Baseline resting-state connectivity was analysed in 40 MCI participants (15 converters, 25 stable). Higher TAF-Net risk was associated with lower within-default-mode connectivity (Spearman *ρ* = - 0.31, p = 0.050), but the association was not specific to the default-mode network: risk correlated with reduced within-network connectivity across multiple association and sensory systems — most strongly the salience/ventral-attention network (*ρ* = -0.39, p = 0.013) — and nine of eleven measures were negatively signed (Figure 6). Consolidating this coherent pattern into a within-network integrity index yielded a stronger association (mean composite *ρ* = -0.41, p = 0.009; first principal component, 37% of shared variance, *ρ* = -0.45, p = 0.003; Figure 6), which also discriminated converters (AUC = 0.71, p = 0.03). Whole-brain global connectivity was only weakly related to risk (*ρ* = -0.20, p = 0.23). An exploratory region-wise analysis localised the association to salience-network hubs (right anterior insula/frontal operculum, dorsal anterior cingulate) and default-mode core regions; 88 of 400 parcels reached nominal significance (about four times chance), though none survived whole-brain FDR at this sample size. Head-motion burden did not account for these associations: TAF-Net risk was unrelated to the number of retained volumes (*ρ* = +0.06, p = 0.72), and partial correlations adjusting for retained volumes were essentially unchanged (integrity composite *ρ* = *−*0.41; within-default-mode *ρ* = *−*0.36). This adjustment is a meaningful test, since retained volumes correlated substantially with the connectivity measures themselves (whole-brain *ρ* = *−*0.52).

**Figure 6.**
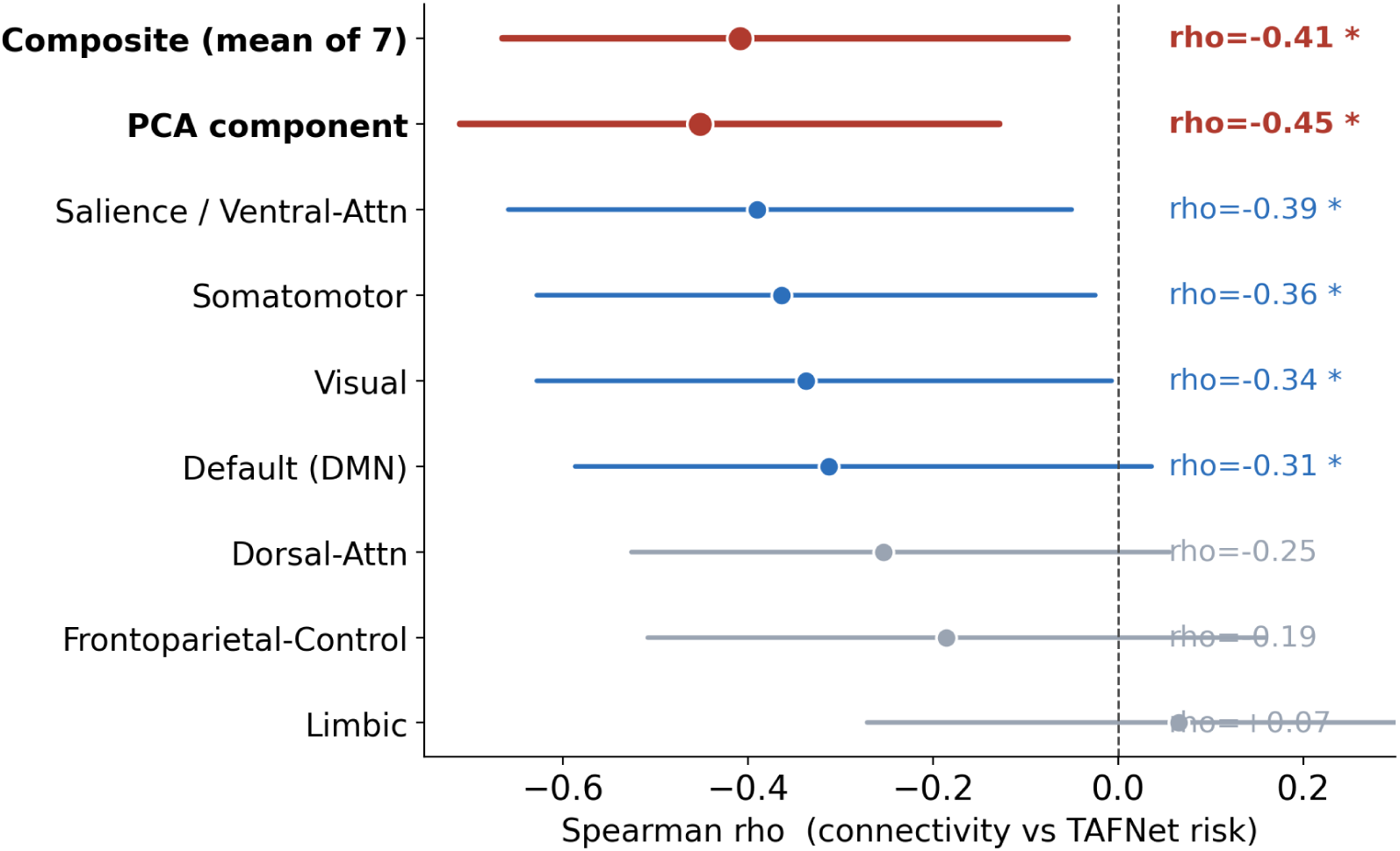
TAF-Net risk tracks within-network connectivity integrity Spearman correlations with resting-state within-network connectivity across the seven Yeo networks and the consolidated integrity indices, with 2000-sample bootstrap confidence intervals; baseline, n = 40. Negative values indicate lower connectivity at higher risk. Red, the consolidated integrity factor.

A second resting-state timepoint was available for only a subset of participants, at highly variable intervals (median 3.5 years, range 1.6–9.6) and predominantly on different scanners. Neither longitudinal change nor endpoint connectivity was significantly related to risk (endpoint *ρ* = -0.05, p = 0.78; risk-change *ρ* = +0.01; time *×* group interaction, n.s.); these analyses were underpowered and are reported as exploratory. Functional grounding is therefore established cross-sectionally, at the baseline timepoint to which the score is anchored.

### 3.7 Calibration and clinical operating characteristics

Discrimination and calibration transferred differently. Although the score ordered participants by risk (Section 3.4), it was poorly calibrated as a probability (Figure 7B). Participants in the lowest tertile of risk received a mean predicted probability of 0.00 yet converted at 15% (Wilson 95% CI 7–31%); in the middle tertile the mean prediction was 0.01 against 15% observed (6–30%); only the highest tertile approached its predicted value (0.46 predicted, 44% observed, 29–61%). The Brier score was 0.197, against a no-skill value of 0.186 obtained by assigning every participant the observed base rate of 24.8%. As a probability estimate the transferred score therefore carried no information beyond the base rate, while retaining substantial discriminative ordering (AUC = 0.72).

**Figure 7.**
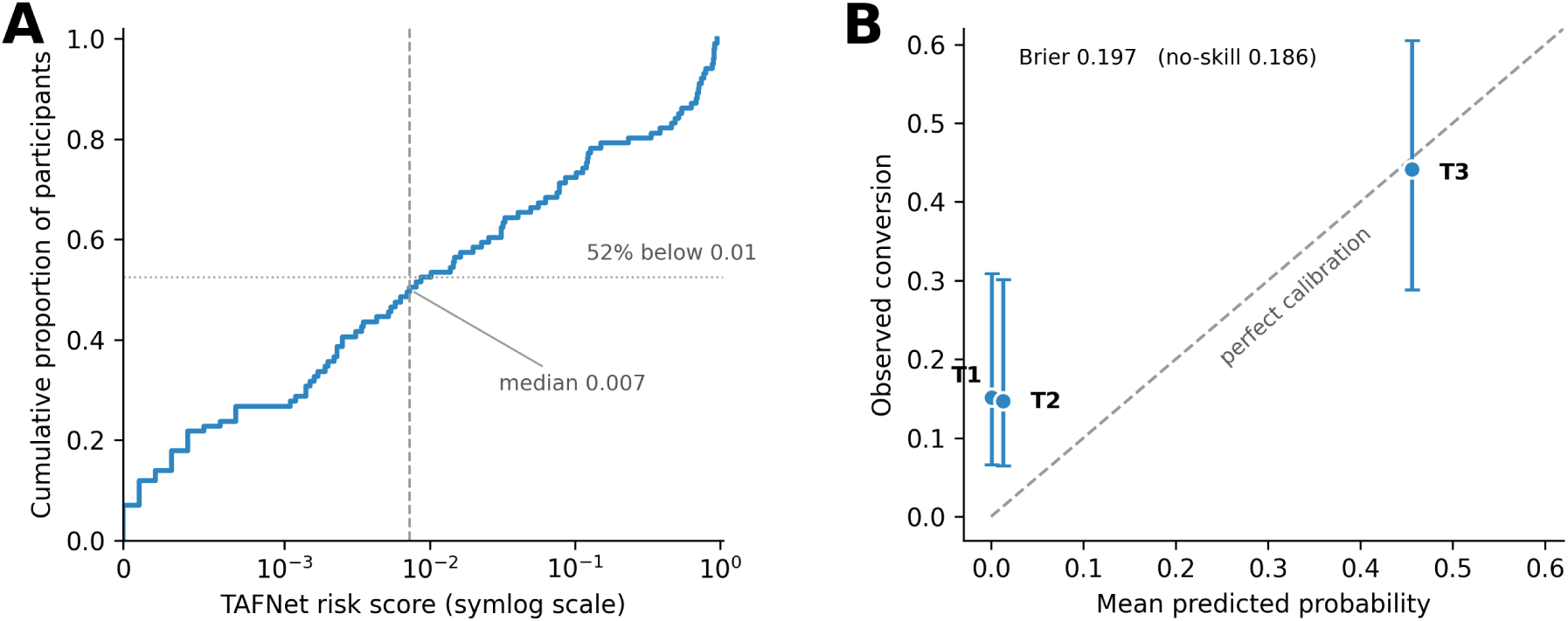
Distribution and calibration of the transferred risk score (A) Distribution of the transferred risk score: empirical cumulative distribution of the participant-level TAF-Net risk (mean across eligible pairs) on a symmetric-log axis; the dashed line marks the median (0.007) and the dotted line the proportion scoring below 0.01 (52%). (B) Calibration by tertile of risk: observed conversion against mean predicted probability for the lowest (T1), middle (T2) and highest (T3) tertiles, with Wilson 95% intervals; the dashed diagonal is perfect calibration, and points above it indicate that the score under-estimates the observed conversion rate. Brier score and its no-skill reference are shown. n = 101.

**Figure 8.**
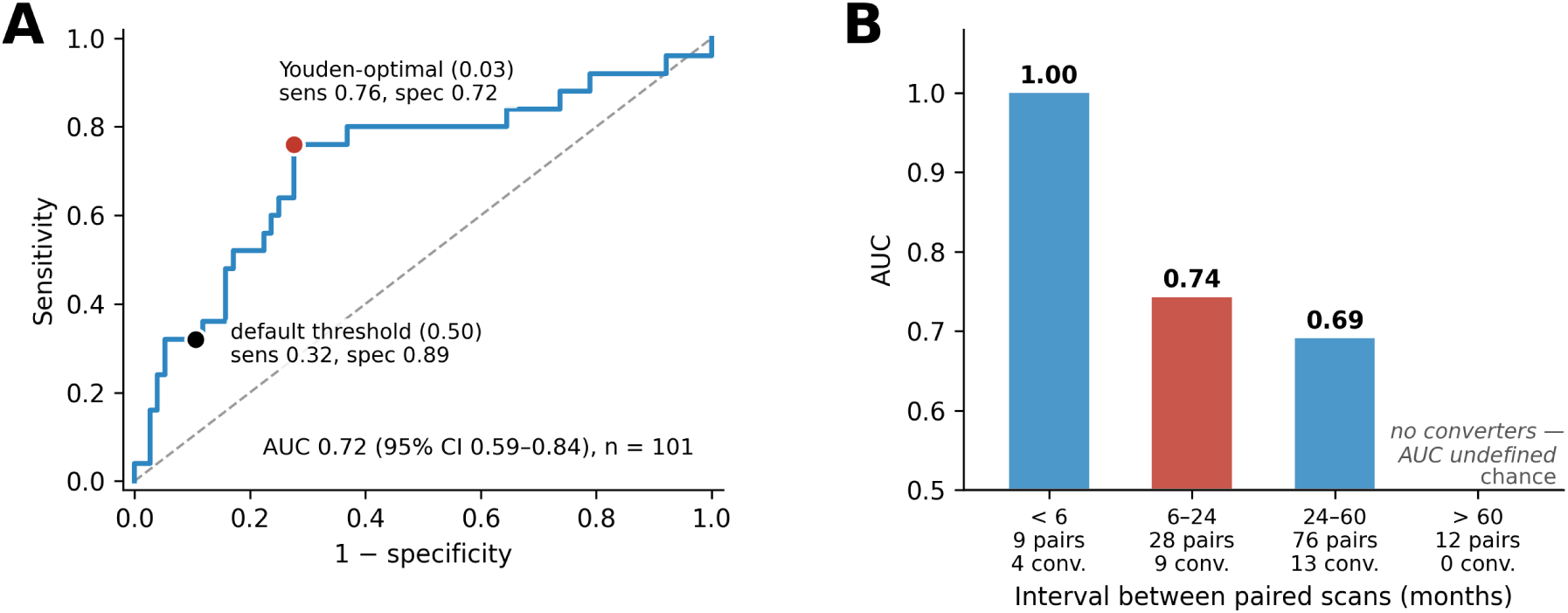
Discrimination in the full cohort and across the scan interval (A) Discrimination in the full cohort: ROC curve for TAF-Net risk in all 101 participants (AUC with bootstrap 95% CI over participants); the black point marks the model’s default threshold of 0.50 and the red point the Youden-optimal threshold of 0.03, each with its sensitivity and specificity. (B) Transfer across the scan interval: AUC at the level of individual scan pairs (n = 125), by interval between the paired scans, with the number of pairs and converters in each stratum; the 6–24-month stratum (red) is the range on which TAF-Net was trained. The under-6-month stratum is too small to be informative, and the *>*60-month stratum contains no converters, so no AUC is defined.

The miscalibration was one-directional: the score under-called conversion. Of the 25 converters, five received scores below 0.01 and only eight exceeded the default threshold of 0.5 — the same number as among the 76 stable participants — so a low score is not reassuring in this cohort even though the ranking is informative. A simple logistic recalibration of the score, fitted in-sample, reduced the Brier score to 0.162, indicating that the deficit lies in the mapping from score to probability rather than in the ordering itself. Because the grounding analyses of Sections 3.2–3.6 used rank-based statistics, they depend only on that ordering and are unaffected by the miscalibration.

## 4. Discussion

Applying an ADNI-trained deep-learning conversion model to an independent OASIS-3 MCI cohort without retraining, we found that its continuous risk scores were grounded in the structural, functional, and cognitive hallmarks of Alzheimer’s disease. Higher risk tracked atrophy that was anatomically specific to AD-signature medial-temporal and temporoparietal regions and absent in AD-spared cortex; discriminated converters comparably to native structural atrophy; tracked the multi-year rate of cognitive decline; and was associated with reduced within-network functional-connectivity integrity at baseline. Across three independent measurement modalities, the predictions align with the neurobiology and clinical course of AD rather than with dataset-specific or non-specific image features.

The anatomical specificity is the strongest evidence for grounding. After adjustment for global atrophy, risk remained coupled to atrophy selectively in entorhinal cortex, amygdala, hippocampus, and inferior and middle temporal cortex — the earliest sites of tau-related neurodegeneration [5, 6] — while spared sensorimotor and visual regions showed no coupling (Steiger Z = -3.23). The model saw only raw longitudinal T1w images, so recovering this canonical topography indicates that it has learned features of the AD atrophy signature rather than global or incidental shrinkage [7, 36].

In external validation, the transferred model discriminated converters in the full cohort (AUC = 0.72) and, in the imaging subsample (AUC = 0.71), at a level indistinguishable from native FreeSurfer atrophy, and the two were strongly concordant; in a joint model, atrophy accounted for the prognostic signal. This is best read not as a weakness but as further evidence of grounding: the model re-expresses, from images alone, the neurodegenerative substrate that regional atrophy captures. Clinically, a single automated score from routine structural MRI reproduced a risk ordering that would otherwise require longitudinal FreeSurfer morphometry, and this ordering transferred across scanners and cohorts (Section 3.4); whether a black-box score should be preferred to the interpretable morphometry it approximates is a separate question [37].

The dissociation between discrimination and calibration is itself informative. Rank ordering transferred across cohorts while absolute risk did not. This cannot be attributed to a difference in base rate — converters made up 26% of the ADNI training set and 25% of this cohort — and more plausibly reflects the other shifts between the two settings: scan intervals largely outside the training range, different scanners and acquisition protocols, and a conversion criterion defined on global CDR rather than clinical diagnosis. A score of this kind is usable for stratifying an independent cohort, and for selecting the upper tail for enrichment, but not — without recalibration — for communicating an individual’s probability of conversion. The gap between the default threshold and the Youden-optimal one in this cohort (0.50 versus 0.03) makes the same point operationally: a transferred model needs its operating point re-derived, not inherited.

The functional findings add a complementary pillar. Higher risk was associated with reduced within-network connectivity integrity, distributed rather than confined to the default-mode network and led by the salience/ventral-attention network, with regional peaks in the anterior insula and dorsal anterior cingulate. This is consistent with the network-degeneration view of AD, in which large-scale association networks lose intrinsic coherence as pathology spreads, and with evidence that default-mode and salience systems are affected early [38, 39, 40, 41]. Meta-analytic evidence indicates that disruption extends beyond the default-mode network to the salience system; in established AD this appears as increased connectivity of the right anterior insula, whereas no consistent salience change has been reported in MCI [42]. The present finding — lower within-salience coherence at higher risk, peaking in the same right anterior insula — was obtained within a risk-stratified MCI sample rather than a case–control contrast. Whether reduced coherence in the prodromal stage precedes the hyperconnectivity reported in dementia warrants direct longitudinal comparison. That the effect was strongest for a consolidated integrity factor and weak for whole-brain connectivity suggests risk relates to the coherence of functional systems rather than to global connectivity level.

## 5. Limitations and future directions

Several limitations temper these conclusions. The structural and functional analyses rest on modest samples (n = 41 and n = 40) determined by imaging completeness, limiting power; no individual functional parcel survived whole-brain correction, and the functional results are therefore reported as a coherent network-level pattern rather than region-specific claims. The functional grounding is also cross-sectional: a longitudinal connectivity analysis was underpowered, with few converters, variable inter-scan intervals, and scanner changes that confound within-subject change.

Confounding is a second concern. The structural analyses were adjusted for global atrophy and head size but not for age, and the sex distribution differed between groups; although age did not differ between converters and stable participants, residual confounding cannot be excluded. Global-signal regression can introduce distance-dependent artifact [27], and the distributed within-network pattern in Section 3.6 should be read with that trade-off in mind.

Finally, the transfer itself has limits. The transferred score was poorly calibrated in this cohort, so the risk values it reports should not be read as probabilities without recalibration; only its ordering is supported here. Most participants also fall outside the 6–24-month scan interval on which the model was trained, and although discrimination degraded only modestly across that shift (Section 3.4), the generality of the transfer beyond this range remains only partly tested. This is also a single external cohort, and grounding against molecular markers — amyloid and tau PET — remains to be established.

These limitations point to clear future directions. First, grounding should be extended to molecular pathology by testing TAF-Net risk against amyloid and tau PET, establishing whether the score tracks the proteinopathies that initiate the disease rather than downstream atrophy alone. Second, the model’s internal spatial attention could be compared directly against the regional atrophy map reported here, converting the present output-level grounding into mechanism-level interpretability. Third, the functional analysis should be repeated in larger, protocol-matched, multi-site samples with statistical harmonisation (e.g., ComBat) [43, 44] and denser longitudinal sampling, so that within-subject connectivity change can be estimated without scanner confounding. Fourth, replication across additional independent cohorts and, ultimately, prospective evaluation in clinical or trial settings will be needed to establish generalisability and real-world utility. More broadly, the multi-modal grounding framework used here — testing a transferred model against independent structural, functional, and cognitive markers — offers a template for validating imaging-based AI biomarkers beyond discrimination metrics alone.

## 6. Conclusion

A deep-learning conversion model trained on one cohort produced risk scores that, applied without retraining to an independent cohort, were grounded in the structural, functional, and cognitive signatures of Alzheimer’s disease. Higher risk selectively tracked AD-signature atrophy, discriminated converters comparably to native morphometry, followed the multi-year course of cognitive decline, and reflected reduced functional-network integrity. These convergent, cross-modal associations indicate that the model’s predictions reflect genuine AD neurodegeneration and generalise across scanners and cohorts, supporting its construct validity as an imaging biomarker while motivating validation against molecular pathology and prospective clinical evaluation.

## Data Availability

The OASIS-3 data analysed in this study are available from the Open Access Series of Imaging Studies (https://www.oasis-brains.org) under its data-use agreement. Derived per-participant measures and analysis code are available from the authors on reasonable request.

## Ethics statement

OASIS-3 data were collected under protocols approved by the Washington University in St. Louis Institutional Review Board with written informed consent from all participants, and were accessed under the OASIS-3 data-use agreement. This secondary analysis used de-identified data only.

## CRediT author contributions

Sara Fin: Conceptualization, Methodology, Formal analysis, Investigation, Data curation, Visualization, Writing – original draft. Alireza Moayedikia: Conceptualization, Methodology, Software, Supervision, Writing – review and editing.

## Acknowledgements

Data were provided in part by OASIS-3: Longitudinal Multimodal Neuroimaging: Principal Investigators: T. Benzinger, D. Marcus, J. Morris; NIH P30 AG066444, P50 AG00561, P30 NS09857781, P01 AG026276, P01 AG003991, R01 AG043434, UL1 TR000448, R01 EB009352.

## Data and code availability

OASIS-3 data are available from https://www.oasis-brains.org under its data-use agreement. Analysis code is available from the authors on reasonable request.

